# Predominance of antibody-resistant SARS-CoV-2 variants in vaccine breakthrough cases from the San Francisco Bay Area, California

**DOI:** 10.1101/2021.08.19.21262139

**Authors:** Venice Servellita, Alicia Sotomayor-Gonzalez, Amelia S. Gliwa, Erika Torres, Noah Brazer, Alicia Zhou, Katherine T. Hernandez, Maddie Sankaran, Baolin Wang, Daniel Wong, Candace Wang, Yueyuan Zhang, Kevin R Reyes, Dustin Glasner, Xianding Deng, Jessica Streithorst, Steve Miller, Edwin Frias, Mary Rodgers, Gavin Cloherty, John Hackett, Susan Philip, Scott Topper, Darpun Sachdev, Charles Y. Chiu

## Abstract

Associations between vaccine breakthrough cases and infection by SARS coronavirus 2 (SARS-CoV-2) variants have remained largely unexplored. Here we analyzed SARS-CoV-2 whole-genome sequences and viral loads from 1,373 persons with COVID-19 from the San Francisco Bay Area from February 1 to June 30, 2021, of which 125 (9.1%) were vaccine breakthrough infections. Fully vaccinated were more likely than unvaccinated persons to be infected by variants carrying mutations associated with decreased antibody neutralization (L452R, L452Q, E484K, and/or F490S) (78% versus 48%, p = 1.96e-08), but not by those associated with increased infectivity only (N501Y) (85% versus 77%, p = 0.092). Differences in viral loads were non-significant between unvaccinated and fully vaccinated persons overall (p = 0.99) and according to lineage (p = 0.09 – 0.78). Viral loads were significantly higher in symptomatic as compared to asymptomatic vaccine breakthrough cases (p < 0.0001), and symptomatic vaccine breakthrough infections had similar viral loads to unvaccinated infections (p = 0.64). In 5 cases with available longitudinal samples for serologic analyses, vaccine breakthrough infections were found to be associated with low or undetectable neutralizing antibody levels attributable to immunocompromised state or infection by an antibody-resistant lineage. Taken together, our results suggest that vaccine breakthrough infecions are overrepresnted by circulating antibody-resistant SARS-CoV-2 variants, and that symptomatic breakthrough infections may potentially transmit COVID-19 as efficiently as unvaccinated infections, regardless of the infecting lineage.

## Introduction

Vaccines targeting the severe acute respiratory syndrome coronavirus 2 (SARS-CoV-2) have been highly effective in preventing symptomatic illness and in reducing hospitalizations and deaths from coronavirus disease 2019 (COVID-19) ^1-8^. Prior studies have also suggested that vaccination may reduce viral loads in persons with breakthrough SARS-CoV-2 infection who have received at least one dose ^7,9^, thus potentially decreasing infectiousness and mitigating transmission. However, most of these studies were done prior to the emergence of “antibody-resistant” SARS-CoV-2 variants of concern / variants of interest (VOCs/VOIs) carrying key mutations that have been shown to decrease antibody (Ab) neutralization (L452R/Q, E484K/Q, and/or F490S), including the Beta (B.1.351), Gamma (P.1), Delta (B.1.617.2), Epsilon (B.1.427/B.1.429), and Lambda (C.37)^10^, but not Alpha (B.1.1.7) variants ^11-13^. Breakthrough infections have been reported in a small proportion of vaccine recipients ^3,14-17^, yet little is known regarding the relative capacity of different variants to escape vaccine-induced immunity and facilitate ongoing spread within highly vaccinated communities.

In San Francisco County, a sharp decline in COVID-19 cases following a 2020-2021 winter outbreak of the Epsilon variant in California ^11,18^ preceded mass vaccination efforts (Figure 1A). From February to June 2021, the number of cases per day continued to gradually decrease, despite a nationwide outbreak from the Alpha variant in the United States ^19^ and the continual introduction of other VOCs/VOIs into the community ^18^. In late June, there was an uptick of delta variant cases presaging a surge of infections from this variant in the county and nationwide ^18^.

**Figure 1.**
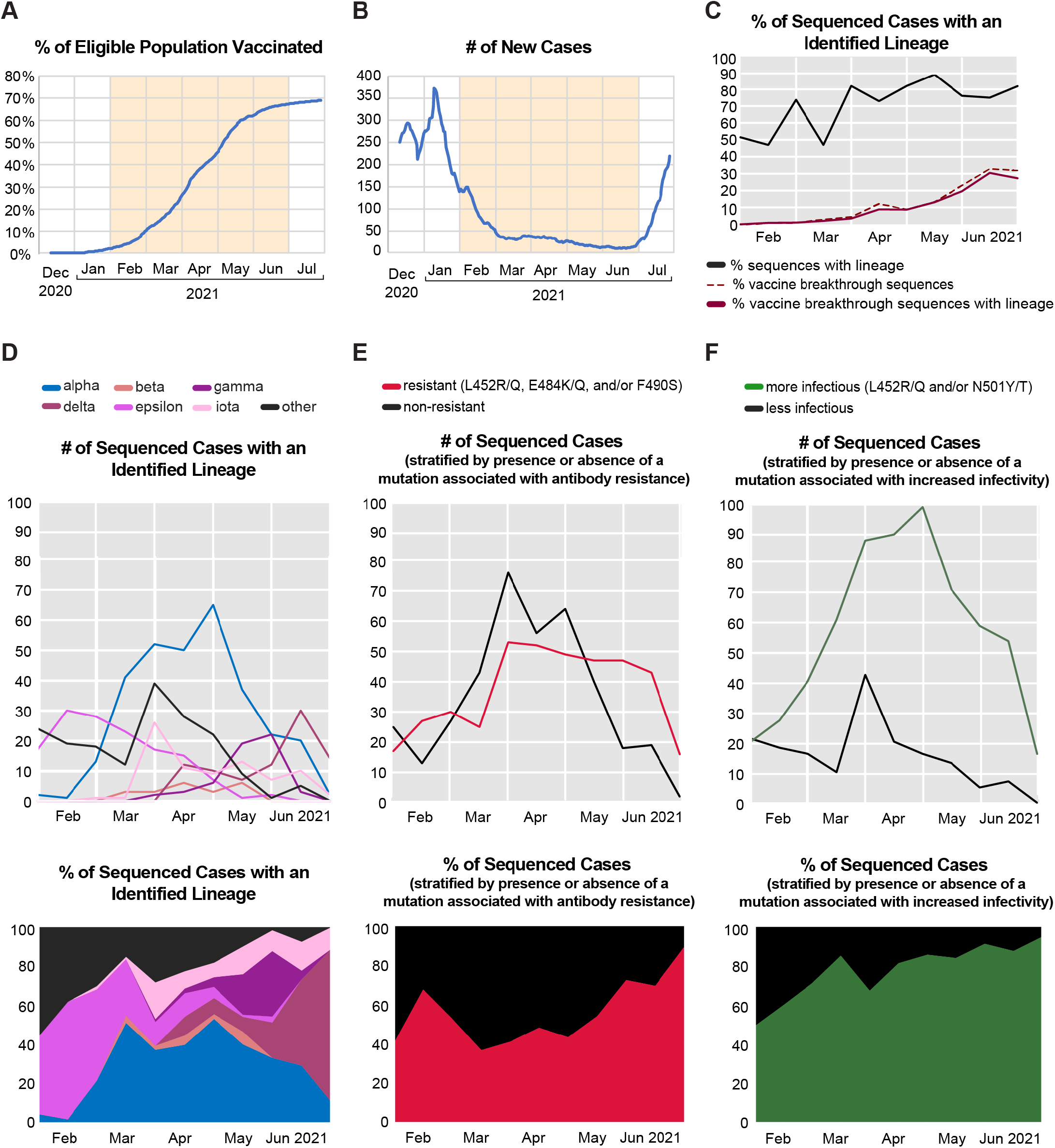
Overview of vaccination and SARS-CoV-2 whole-genome sequencing data from the San Francisco Bay Area. **(A)** Plot showing the percentage of eligible individuals in San Francisco County who had received an FDA-authorized vaccine from the beginning of mass vaccine rollout until June 30, 2021. The peach-colored shaded area denotes the study timeframe for sample collection. **(B)** Plot showing the 7-day rolling average number of new SARS-CoV-2 positive cases in San Francisco County. **(C)** Plot showing the proportions of total vaccine breakthrough sequences (denoted by the dotted line) and vaccine breakthrough sequences with identified lineages, relative to the total number of sequenced cases, and aggregated biweekly. (**D-F)** Plots showing the counts (top) and proportion (bottom) of SARS-CoV-2 positive sequenced cases aggregated biweekly and stratified by **(D)** lineages identified by Pangolin algorithm^36^, **(E)** presence/absence of mutations associated with antibody resistance, and **(F)** presence/absence of mutations associated with increased infectivity.

Here we performed whole-genome sequencing and viral load measurements of nasal swabs in conjunction with retrospective medical chart review from COVID-19 infected persons over a 5-month time period to investigate dynamic longitudinal shifts in the distribution of SARS-CoV-2 variants over time and to identify potential correlates of breakthrough infections in a progressively vaccinated community.

## Results

We performed whole-genome sequencing of available remnant mid-turbinate nasal, nasopharyngeal and/or oropharyngeal (OP) swab samples collected from 1,373 PCR-positive COVID-19 cases from San Francisco County from February 1 to June 30, 2021. During this study period, the percentage of eligible persons vaccinated in the county increased from 2 to 70%, while the 7-day rolling average number of cases per day declined from 150 to 20. (Figure 1A and B). The cohort included COVID-19 patients seen in hospitals and clinics at University of California, San Francisco (UCSF, n=598, 43.6%) and infected persons identified by community testing in San Francisco County performed by a commercial laboratory (Color Genomics, n=775, 56.4%). Using the CDC definition of a vaccine breakthrough infection as a positive SARS-CoV-2 RNA or antigen test ≥14 days after completion of all recommended doses ^20^, 125 (9.1%) of infections in the cohort were vaccine breakthroughs (Table 1), and the percentage of sequenced cases that were vaccine breakthroughs increased from 0% to 31.8% from February to June **(Figure 1C)**. Among the viruses sequenced from the 1,373 cases, 69% (945 of 1,373) were unambiguously assigned to a SARS-CoV-2 lineage (Table 1).

**Table 1.**
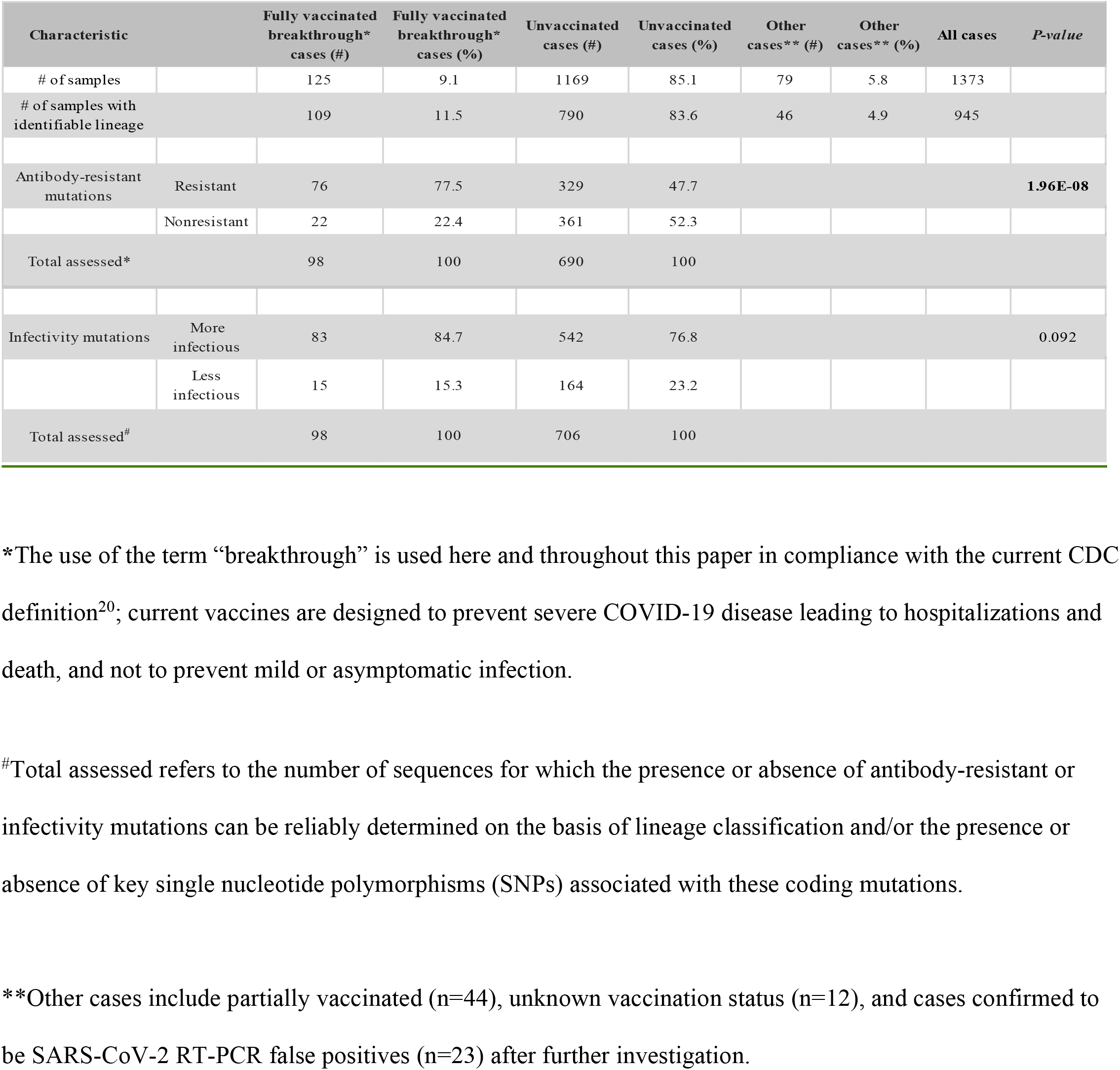
Characteristics of vaccine breakthrough and unvaccinated cases.

| Characteristic |  | Fully vaccinated<br>breakthrough*<br>cases (#) | Fully vaccinated<br>breakthrough*<br>cases (%) | Unvaccinated<br>cases (#) | Unvaccinated<br>cases (%) | Other<br>cases** (#) | Other<br>cases** (%) | All cases | P-value |
| --- | --- | --- | --- | --- | --- | --- | --- | --- | --- |
| # of samples |  | 125 | 9.1 | 1169 | 85.1 | 79 | 5.8 | 1373 |  |
| # of samples with<br>identifiable lineage |  | 109 | 11.5 | 790 | 83.6 | 46 | 4.9 | 945 |  |
| Antibody-resistant<br>mutations | Resistant | 76 | 77.5 | 329 | 47.7 |  |  |  | **1.96E-08** |
|  | Nonresistant | 22 | 22.4 | 361 | 52.3 |  |  |  |  |
| Total assessed* |  | 98 | 100 | 690 | 100 |  |  |  |  |
| Infectivity mutations | More<br>infectious | 83 | 84.7 | 542 | 76.8 |  |  |  | 0.092 |
|  | Less<br>infectious | 15 | 15.3 | 164 | 23.2 |  |  |  |  |
| Total assessed<sup>#</sup> |  | 98 | 100 | 706 | 100 |  |  |  |  |

\*The use of the term “breakthrough” is used here and throughout this paper in compliance with the current CDC definition<sup>20</sup>; current vaccines are designed to prevent severe COVID-19 disease leading to hospitalizations and death, and not to prevent mild or asymptomatic infection.
<sup>#</sup>Total assessed refers to the number of sequences for which the presence or absence of antibody-resistant or infectivity mutations can be reliably determined on the basis of lineage classification and/or the presence or absence of key single nucleotide polymorphisms (SNPs) associated with these coding mutations.
\*\*Other cases include partially vaccinated (n=44), unknown vaccination status (n=12), and cases confirmed to be SARS-CoV-2 RT-PCR false positives (n=23) after further investigation.

Over the 5-month study period, the proportion of circulating antibody-resistant variants, or those containing the L452R/Q, E484 K/Q, and/or F490S mutations, was 47.7% (329 of 690 sequenced genomes with identifiable mutations) (Table 1), while the proportion of variants with increased infectivity, or those containing the L452R/Q, F490S, and/or N501T/Y mutations, was 76.8% (542 of 706 sequenced genomes with identifiable mutations). The proportion of antibody-resistant variants, calculated by aggregating cases with lineages carrying resistance-associated mutations over 2-week intervals, increased from 40% to 89% (Figure 1E), while the proportion of variants with increased infectivity increased from 49% to 94% (Figure 1F). In unvaccinated cases, most viruses consisted of non-resistant variants (61% and 57% based on community and UCSF testing, respectively) (Figure 2A, right), in contrast to vaccinated cases, for which the proportions of non-resistant variants dropped to 34% and 20%, respectively (Figure 2A, left). To account for temporal trends in the circulating variants and the percentage of vaccinated population, we calculated the proportion of antibody-resistant variants in vaccinated and unvaccinated cases per month. Interestingly, we found statistically significant higher proportions of antibody-resistant variants in post-vaccinated cases in April, May, and June 2021. Of note, Alpha was the predominant variant of concern in California in April and May 2021^40^. We found no significant differences in February and March 2021, albeit the number of vaccinated breakthrough cases during these months were very low (Figure 4). Alpha was the only non-resistant variant associated with breakthrough infection in vaccinated cases. Overall, fully vaccinated cases were significantly more likely than unvaccinated cases to be infected by resistant variants (77.6% versus 47.7%, p=1.96e-08) (Figure 2B, top and Table 1), but not by variants associated with increased infectivity (84.7% versus 76.8%, p = 0.092) (Figure 2B, bottom and Table 1). The distribution of variants in immunocompetent and immunocompromised patients was similar (Figure 2A). Infections by the Gamma and Delta variants, which cause more pronounced decreases in Ab neutralization relative to most of the other resistant VOCs ^12,21^ (Figure 2A, left) were increased in fully vaccinated breakthrough infections as compared to unvaccinated infections. In contrast, variant distribution in unvaccinated cases, with Alpha and Epsilon predominant (Figure 2B, right), was similar to estimates of prevalence locally in the community and in the state of California during the study period^11,18^

**Figure 2.**
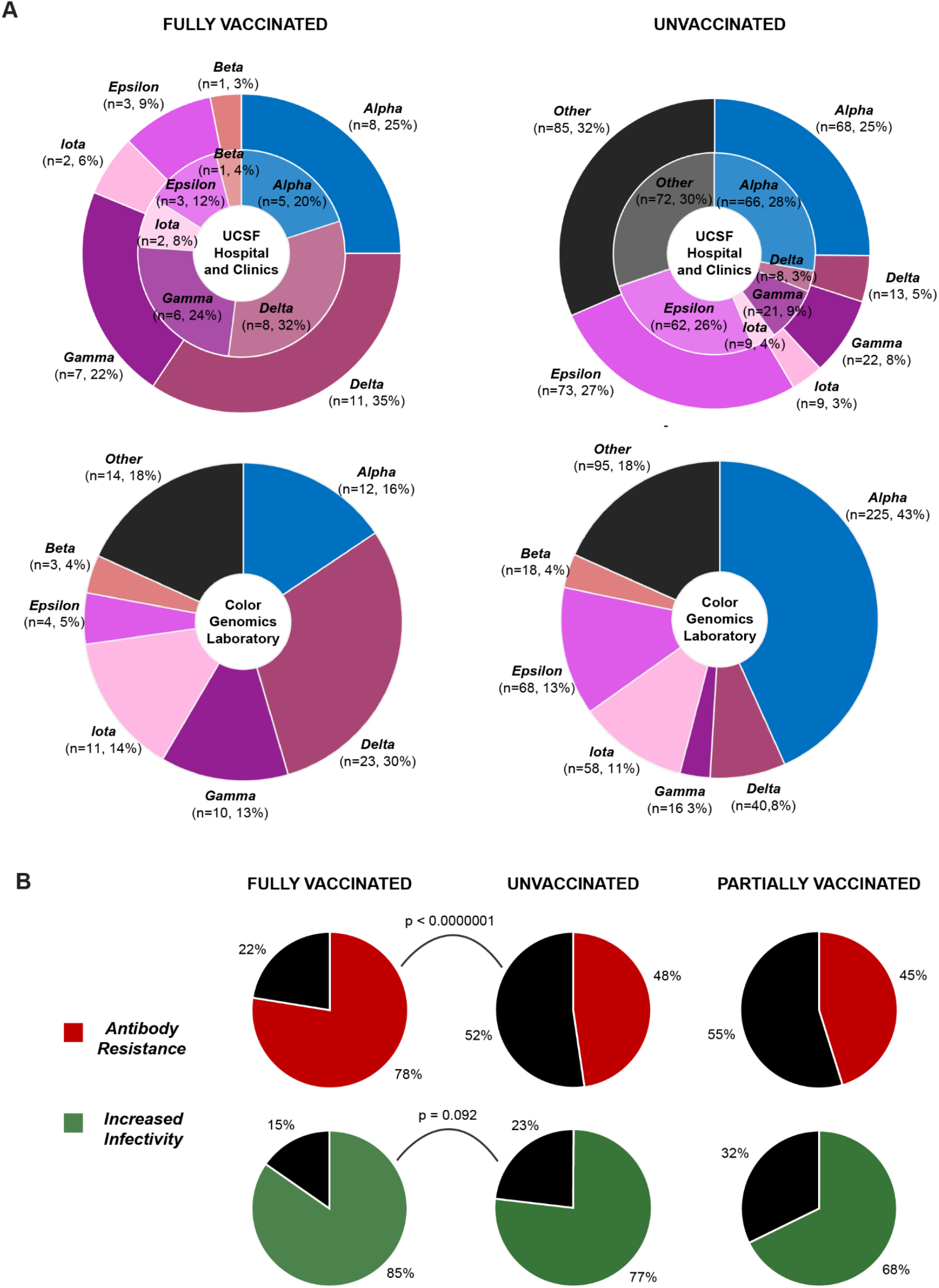
Comparisons of lineage distribution and proportion of mutations associated with antibody resistance and increased infectivity in fully vaccinated, unvaccinated, and partially vaccinated cases. **(A)** Pie charts showing the distribution of SARS-CoV-2 variant lineages in fully vaccinated and unvaccinated cases from UCSF Hospitals and Clinics (top) and from Color Genomics Laboratory (bottom). In the UCSF charts, the inner circles represent the immunocompetent cases, and the outer circles include both immunocompetent and immunocompromised individuals. **(B)** Pie charts showing the proportions of SARS-CoV-2 genomes carrying mutations associated with antibody resistance (top) and increased infectivity (bottom) in fully vaccinated and unvaccinated cases from UCSF Hospitals and Clinics and Color Genomics Laboratory, and partially vaccinated cases from UCSF. Red color indicates the presence of mutations associated with antibody resistance, green indicates the presence of mutations associated with increased infectivity, and black indicates the absence of either mutations.

Viral RNA loads for infected persons in the full patient cohort, 125 of whom were vaccine breakthrough cases, were estimated by comparison of differences in mean cycle threshold (Ct) values using quantitative RT-PCR. There was no difference in viral RNA loads between fully vaccinated breakthrough and unvaccinated cases, either overall (p = 0.99) or according to lineage (p = 0.09 – 0.78) (Figures 3A and 3B). Infections from variants of concern (VOCs/VOIs) had significantly higher viral loads than non-VOC/VOI lineages overall (p=0.017, Δ Ct = 1.2) (Figure 3C). With respect to individual VOCs, higher viral RNA loads were observed for infections by the Gamma (p=0.00076, Δ Ct = 2.5), delta (p=0004, Δ Ct = 2.1), and Epsilon (p=0.047, Δ Ct = 1.2) variants, but not for the Alpha, Beta, and Iota variants (Figure 3D).

**Figure 3.**
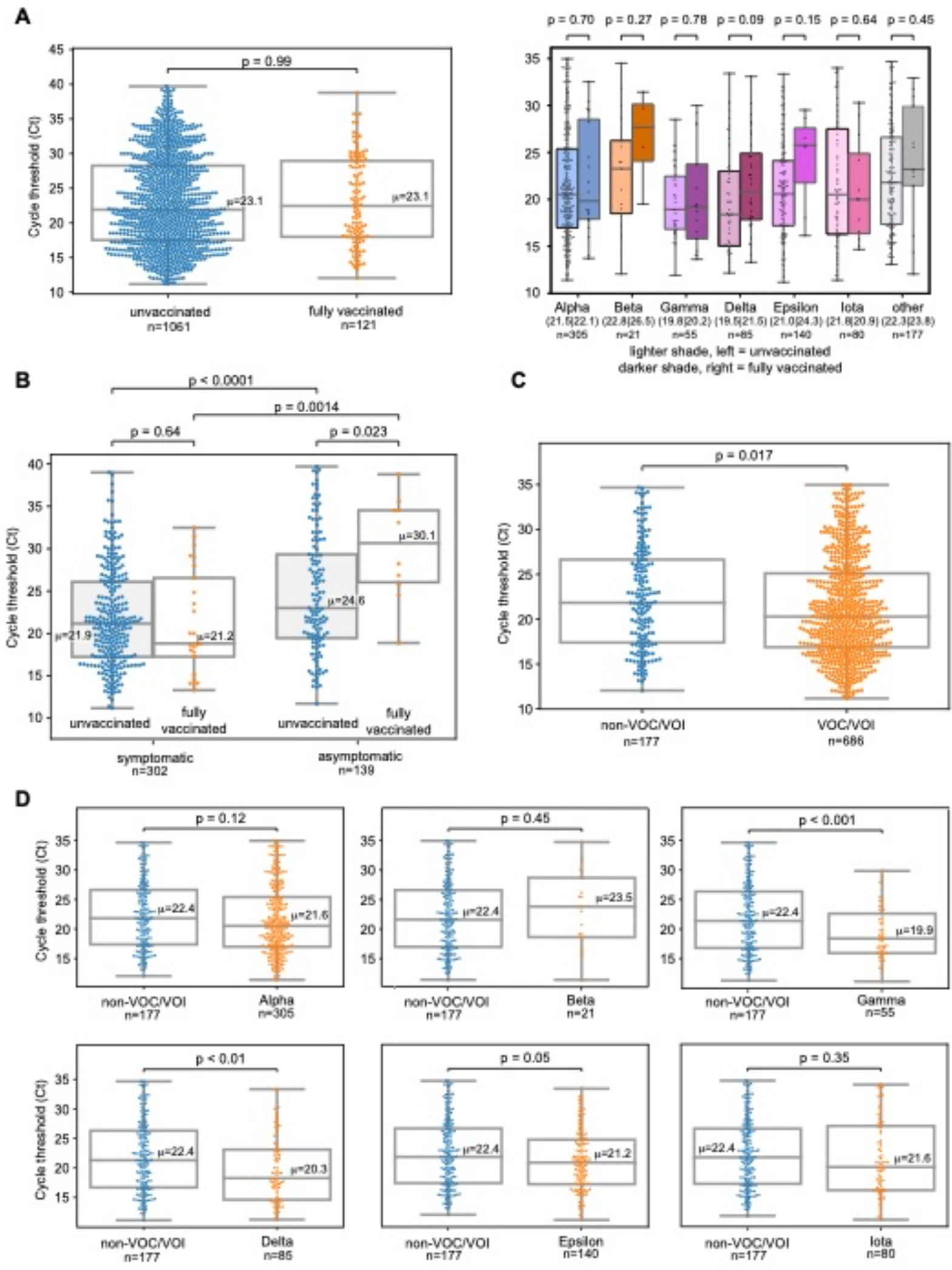
Comparison of viral loads according to clinical symptoms, vaccination status, and SARS-CoV-2 lineage. (A) Grouped box-and-whisker plots and swarm plots showing the differences in mean cycle threshold (Ct) values between vaccinated and unvaccinated cases overall (left) and stratified according to lineages (right). There were no significant differences in viral loads (inversely proportional to the Ct value) in both comparisons. Notably, higher viral loads were observed in Gamma and Delta unvaccinated cases, but the differences were not statistically significant (p > 0.05). (**B)** Box-and-whisker and swarm plots showing the differences in viral loads between symptomatic and asymptomatic cases. Each group was further subdivided into fully vaccinated and unvaccinated cases. Each pairwise comparison was statistically significant except for symptomatic unvaccinated and symptomatic vaccinated. **(C)** Box-and-whisker and swarm plots showing the differences in viral load between lineages identified as VOC/VOI (variant of concern/variant of interest) and other lineages that are not VOCs or VOIs (“non-VOC/VOI”). In this study, identified VOCs/VOIs included Alpha, Beta, Gamma, Delta, Epsilon, and Iota variants, following the World Health Organization (WHO) nomenclature scheme. **(D)** Box- and-whisker and swarm plots showing differences in viral loads between each VOC/VOI and non-VOC/VOI. Blue-colored dots denote non-VOC/VOI data points while orange-colored dots denote VOC/VOI data points. For all box-and-whisker plots, the box outlines denote the interquartile ratio (IQR), the solid line in the box denotes the mean (μ) Ct value, and the whiskers outside the box extend to the minimum and maximum fold enrichment points. For the lineage plots in **3A (right)**, the mean Ct values corresponding to unvaccinated and fully vaccinated cases are provided as μ_unvaccinated_|μ_vaccinated_ in parentheses next to the name. Welch’s t-test was used for significance testing.

**Figure 4.**
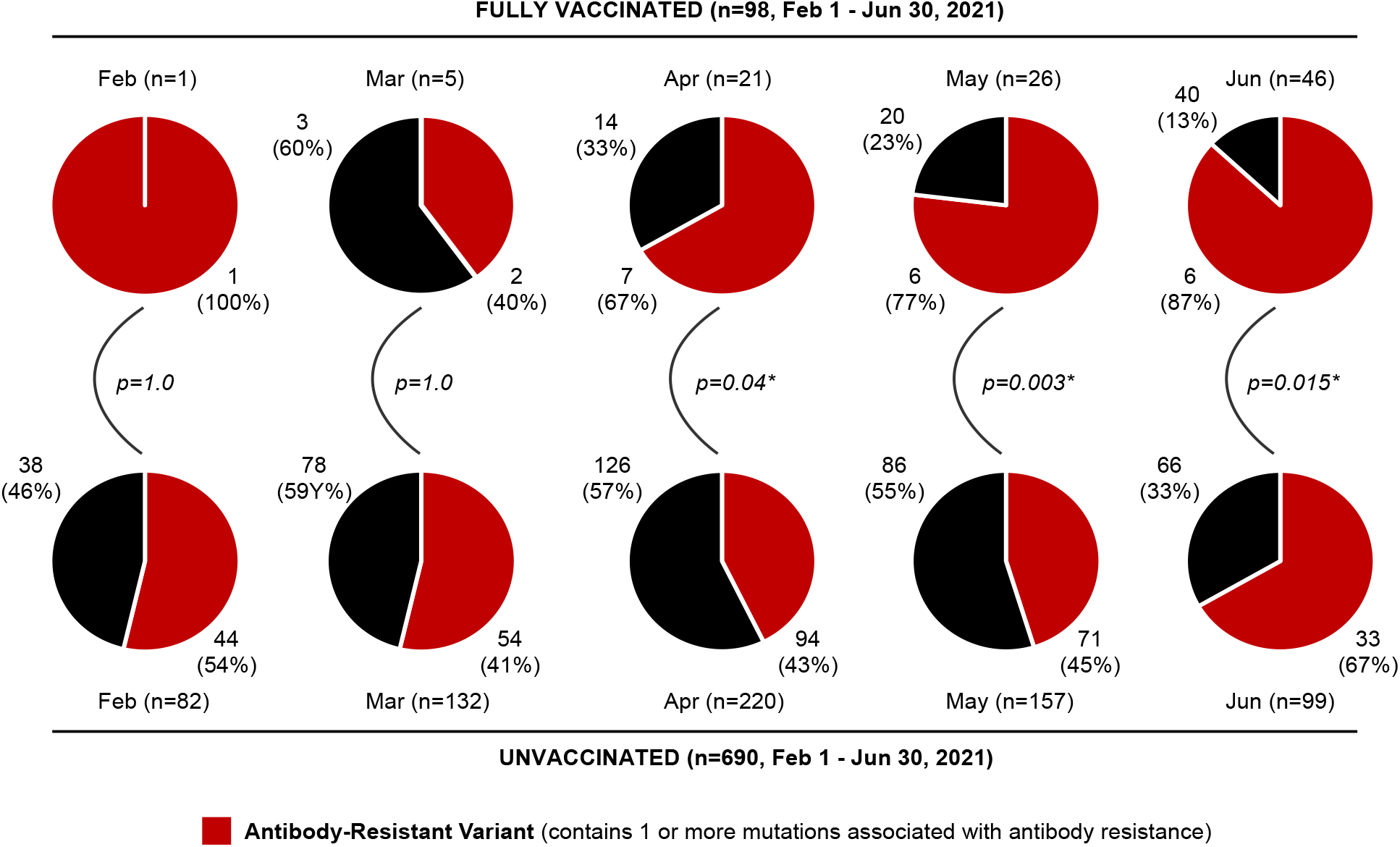
Comparisons of proportion of mutations associated with antibody resistance in fully vaccinated and unvaccinated cases by month. Pie charts showing the monthly proportions of SARS-CoV-2 genomes carrying mutations associated with antibody resistance in fully vaccinated and unvaccinated cases from UCSF Hospitals and Clinics and Color Genomics Laboratory from February 2021 to June 2021. Red color indicates the presence of mutations associated with antibody resistance. Fisher’s exact test method was used to determine the *P* values.

We investigated potential correlations between variant identification, clinical symptomatology, vaccine type, and viral load in vaccine breakthrough infections. Retrospective medical chart review was performed for a subset of patients from UCSF hospitals and clinics with available clinical and demographic data (n=598) (Table 2). Among the 39 breakthrough infections out of 598 in this subset, the average age was 49 years (range 22 to 97), and the majority were women (54%). The median interval from completion of all doses of the vaccine and COVID-19 breakthrough infection was 73.5 days (range 15 to 140). The Pfizer-BioNTech (BNT162b2) COVID-19 mRNA vaccine was administered to 20 (51%) of the vaccine breakthrough patients, while 11 (28%) received the Moderna (mRNA-1273) COVID-19 mRNA vaccine and 4 (10%) received the Johnson & Johnson/Janssen (JnJ) COVID-19 viral vector (adenovirus) vaccine. Nine (23%) of the vaccine breakthrough patients were immunocompromised, while 28 (72%) were identified as symptomatic and 10 (26%) as asymptomatic. Among the symptomatic breakthrough infections, 6 patients (15.4%) were hospitalized for COVID-19 pneumonia, 1 patient (2.6%) required care in the intensive care unit (ICU), and 0 patients (0%) died. Among the unvaccinated infections (n=433), 287 (66%) patients presented with symptoms while 132 (30%) patients reported no symptoms; 51 patients (11.8%) required hospitalization, 27 (6.2%) patients were admitted to the ICU and 5 (1.2%) patients died with COVID-19 reported as the primary cause of death. Among these clinical and demographic variables, only advanced age of >65 years was significantly associated with vaccine breakthrough infections as compared to unvaccinated infections (p = 0.035, odds ratio (OR) 2.36 [95% CI 0.97, 5.45]). Viral RNA loads were significantly higher overall for symptomatic as compared to asymptomatic infections for both vaccine breakthrough (p = 0.0014, Δ Ct = 8.8 [95% CI 4.0 – 13.7]) and unvaccinated (p = 9.8e-05, Δ Ct = 2.8 [95% CI 1.42 – 4.21]) cases (Figure 3B). Differences in RNA viral loads for vaccine breakthroughs as compared to unvaccinated cases were non-significant for symptomatic cases (p = 0.64, Δ Ct = 0.6 [95% CI −2.0 – 3.2]), but were significant for asymptomatic cases (p = 0.023, Δ Ct = −5.4 [95% CI −9.9 – 0.09]), while viral RNA loads of hospitalized patients with COVID-19 did not differ significantly from those of outpatients (Figure S2).

**Table 2.**
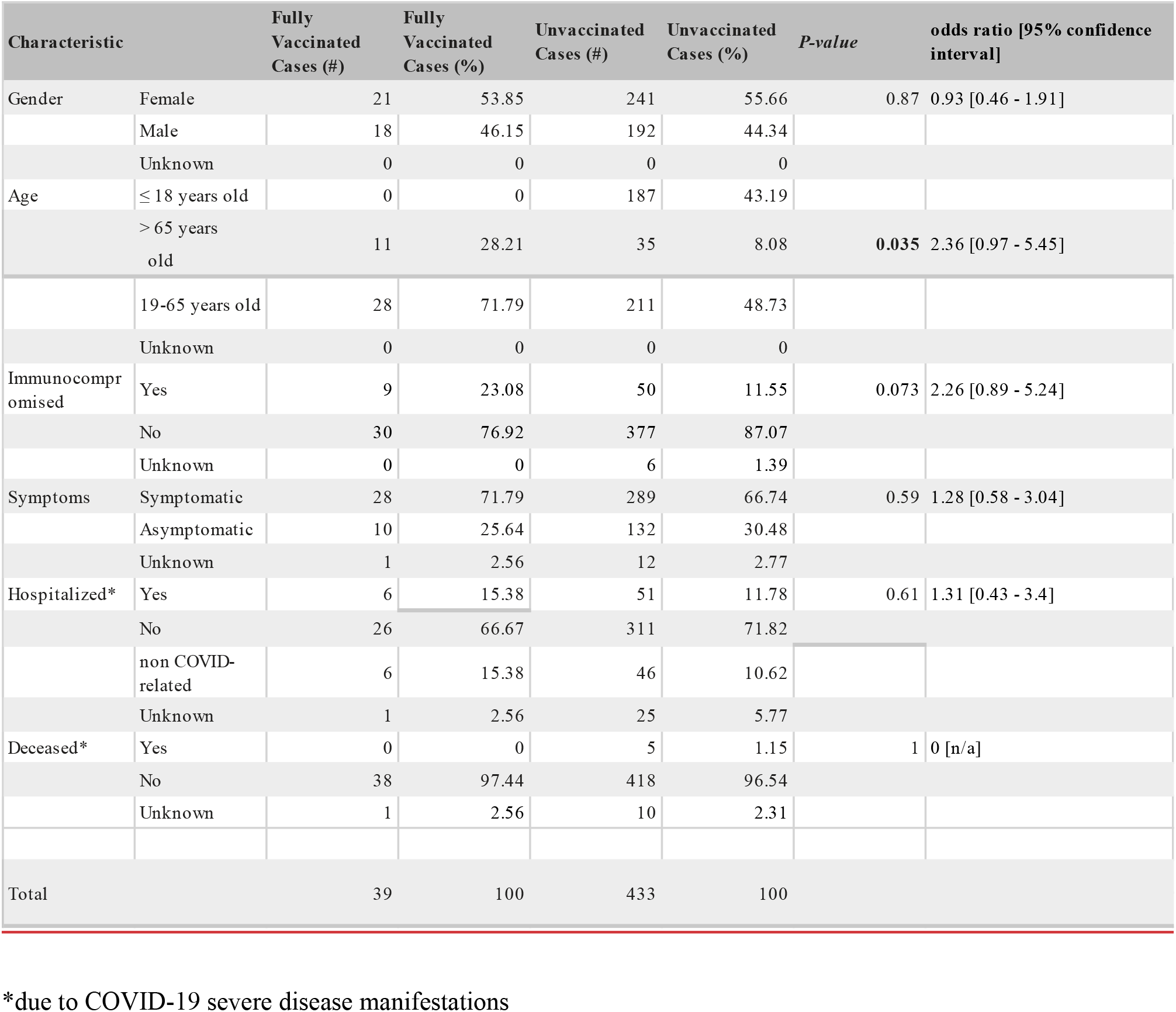
Clinical and demographic characteristics in vaccine breakthrough and unvaccinated cases.

We sought to understand the serologic basis behind some of the vaccine breakthrough infections in the study cohort. Plasma samples were available from 5 of 39 (12.8%) patients with clinical metadata for qualitative testing of nucleoprotein immunoglobulin G (IgG) and spike immunoglobulin M (IgM) levels and neutralizing Ab titers using a cytopathic effect (CPE) endpoint neutralization assay as previously described ^11^. For 4 of 5 patients, longitudinally collected samples were available. Neutralization assays were tested for activity against cultures of a wild-type (early 2020 non-VOC) SARS-CoV-2 virus and the Alpha, Beta, Gamma, Delta, and Epsilon variants (Table 3). In 3 of 5 cases, patients failed to mount detectable qualitative and neutralizing Ab responses to the vaccine, likely because they were immunocompromised (Table 3). The interpretation was indeterminate for one case (Table 3, “P3”), as only samples at day 11 post-breakthrough or later were available, by which time the patient had likely generated a robust antibody response to the breakthrough infection. The remaining case (Table 3, “P4”) was a *bona fide* vaccine breakthrough case in an immunocompetent patient who had received the JnJ vaccine and had also been previously infected with COVID-19 prior to vaccination. This patient was negative for detection of qualitative nucleoprotein IgG and spike protein IgM Ab from plasma 2 days after testing SARS-CoV-2 positive from NP swab by RT-PCR; however, strong positivity for spike protein IgG Ab and high neutralizing Ab titers against WT D614G virus suggests a robust antibody response to vaccination. Levels of neutralizing Ab were lowest for the Beta, Delta, and Epsilon variants, consistent with the patient’s breakthrough infection by Delta.

**Table 3.** Qualitative and neutralizing antibody studies in vaccine breakthrough patients.

| Patient # | Infecting lineage | Vaccine received | Imm? | Clinical presentation of breakthrough | Plasma sample collection day (relative to day of first positive respiratory swab PCR) | SARS-CoV-2 IgG Ab (spike RBD) | SARS-CoV-2 IgG Ab (nucleoprotein) | SARS-CoV-2 IgM Ab (spike) | Antibody neutralization |  |  |  |  |  | Interpretation (reason for vaccine failure) |
| --- | --- | --- | --- | --- | --- | --- | --- | --- | --- | --- | --- | --- | --- | --- | --- |
|  |  |  |  |  |  |  |  |  | D614G (WT) | B.1.1.7 (alpha) | B.1.351 (beta) | P.1 (gamma) | B.1.617.2 (delta) | B.1.429 (epsilon) |  |
| P1 | non-VOC (B.1) | Pfizer/BioNTech BNT162b2 | yes | hospitalized with COVID-19 pneumonia | -36 days | - (0.0) | - (1.0) | - (0.02) | <100 | <100 | <100 | <100 | <100 | <100 | no Ab response to vaccine, likely due to immunocompromised state |
|  |  |  |  |  | +9 days | + (157.62) | - (0.41) | + (1.58) | <100 | <100 | <100 | <100 | <100 | <100 |  |
|  |  |  |  |  | +17 days | + (14258.34) | + (4.27) | + (17.11) | 9051 | 898 | 356 | 566 | 898 | 713 |  |
|  |  |  |  |  | +28 days | + (52626.99) | + (8.12) | + (5.41) | 9051 | 898 | 224 | 449 | 566 | 1425 |  |
| P2 | B.1.1.7 (alpha) | Pfizer/BioNTech BNT162b2 | yes | hospitalized with COVID-19 pneumonia | +6 days | - (55.3) | - (0.23) | - (0.01) | <100 | <100 | <100 | <100 | <100 | <100 | no Ab response to vaccine, likely due to immunocompromised state |
|  |  |  |  |  | +9 days | - (41.9) | - (0.26) | - (0.01) | <100 | <100 | <100 | <100 | <100 | <100 |  |
| P3 | B.1.1.7 (alpha) | Moderna mRNA-1273 | yes | hospitalized with COVID-19 pneumonia | +11 days | + (13889.68) | + (4.94) | + (58.97) | 11404 | 11404 | 2263 | 11404 | 3592 | 1796 | indeterminate (no pre-breakthrough sample available) |
|  |  |  |  |  | +18 days | + (23622.01) | + (5.02) | + (45.66) | 4525 | 4525 | 2263 | 4525 | 1425 | 356 |  |
| P4* | B.1.617.2 (delta) | Johnson and Johnson JNJ-78436735. | no | asymptomatic | +2 days | + (13889.68) | - (0.88) | - (0.73) | 11404 | 5702 | 898 | 11404 | 2851 | 2263 | failure to mount adequate Ab response to vaccine; decreased Ab response to delta variant |
| P5 | P.1 (gamma) | Moderna mRNA-1273 | yes | symptomatic (fever, malaise, and myalgia) | +3 days | - (0.0) | - (0.04) | - (0.03) | <100 | <100 | <100 | <100 | <100 | <100 | no Ab response to vaccine, likely due to immunocompromised state |
|  |  |  |  |  | +5 days | - (0.0) | - (0.03) | - (0.03) | <100 | <100 | <100 | <100 | <100 | <100 |  |

\*Patient had a prior infection from COVID-19
Abbreviations: PCR, polymerase chain reaction; RBD, receptor binding domain; IgG, immunoglobulin G; IgM, immunoglobulin M; Ab, antibody;
WT, wild-type

## Discussion

Here we used variant identification by SARS-CoV-2 whole-genome sequencing, quantitative viral load analysis, and antibody studies along with retrospective medical chart review to compare vaccine breakthrough (n=125, 9.1%) and unvaccinated (n=1169, 85.1%) cases in both community and hospitalized settings from northern California. Prior reports have shown that the distribution of VOCs/VOIs in breakthrough cases generally reflect the estimated community prevalence in the unvaccinated population ^14,16,17,22-24^. These reports, however, investigated breakthrough cases over a limited timeframe during which only a single predominant lineage was typically circulating. The current study spanned 5 months as the study population became progressively vaccinated from 2% to >70% while undergoing 3 successive surges of infection from the Epsilon (Feb - Mar 2021) ^11^, Alpha (Mar 2021 - June 2021) ^19^, and Delta (June 2021) variants^18^. In contrast to previous studies, we found that vaccine breakthrough infections are more likely to be caused by immunity-evading variants as compared to unvaccinated infections. These findings are largely attributed to the observed decreased proportion of vaccine breakthrough infections from the Alpha variant, despite its documented higher infectivity relative to all VOCs except Delta and Gamma^11,25-27^. Decreased Alpha infections are consistent with the higher effectiveness of available SARS-CoV-2 vaccines against Alpha relative to other VOCs ^2,15^, most of which exhibit higher resistance to neutralizing antibodies than Alpha ^12,13,15^. The predominance of immune-evading variants among post-vaccination cases indicates selective pressure for immune-resistant variants locally over time in the vaccinated population concurrent with ongoing viral circulation in the community. In particular, the Delta variant, which is the predominant circulating lineage in the United States as of July 2021, has been shown to be more resistant to vaccine-induced immunity as well as being more infectious than Alpha ^13,15,21,25^.

Among demographic and clinical factors associated with vaccine breakthrough infection, we only identified a significant association with age, consistent with the prioritized rollout of the vaccine in the elderly population ^2^. Several studies have demonstrated that the vaccine remains highly effective against preventing symptomatic breakthrough infections resulting in serious illness leading to hospitalization and/or death ^1-8^. Our findings are consistent with these other studies, as there were fewer hospital admissions and no deaths in vaccinated patients as compared to unvaccinated patients, though these differences were not statistically significant due to low case numbers.

We also found that differences in viral RNA loads (as estimated using Ct values) between vaccine breakthrough and unvaccinated infections were non-significant (p = 0.99), regardless of lineage. A previous study of a community outbreak of Delta infections in the state of Massachusetts also found that viral loads were similar for both vaccinated and unvaccinated persons with COVID-19 ^28^. These findings likely formed the basis for revised indoor mask guidance in July 2021 from the United States Centers for Disease Control and Prevention ^29^. Our results show that comparably high viral loads in vaccine breakthrough infections are not confined to Delta alone; indeed, the highest viral loads were observed from Gamma. Notably, as shown in Figure 3B, viral RNA loads in symptomatic vaccine breakthrough cases were approximately 445X higher as compared to asymptomatic cases (p = 0.0014, Δ Ct = 8.8), and similar to those in unvaccinated cases (p = 0.64, Δ Ct = 0.64). However, significantly lower viral RNA loads were observed in asymptomatic breakthrough cases as compared to unvaccinated cases (p=0.0023). Taken together, these data suggest that symptomatic breakthrough cases are likely as infectious as symptomatic unvaccinated cases, and thus may contribute to ongoing SARS-CoV-2 transmission, even in a highly vaccinated community. These findings thus reinforce the importance of mask wearing recommendations in symptomatic persons to control community spread, regardless of vaccination status ^29^. These also suggest that asymptomatic transmission of breakthrough cases may be less efficient given the lower viral loads. Contact tracing investigation of vaccine breakthroughs is likely needed to ascertain the role, if any, of asymptomatic transmission in vaccinated persons to SARS-CoV-2 spread.

Our antibody analyses, although performed on a small number of cases (n=5), show that vaccine breakthrough cases are generally associated with low or undetectable qualitative and neutralizing antibody levels in response to vaccination. These findings are consistent with studies that have correlating high antibody levels with vaccine efficacy^30^. We identified 3 cases of breakthrough infection from Alpha, all in immunocompromised patients who failed to mount detectable levels of neutralizing antibody to both wild-type and VOC SARS-CoV-2 lineages. We also reported 1 case of Delta breakthrough infection in a patient who had contracted COVID-19 in 2020 and who had also received the JnJ vaccine. This case, although singular, demonstrates the likely inadequacy of convalescent antibodies generated from prior infection in protecting against future infection, especially against emerging antibody-resistant VOCs, and the reduced effectiveness of the JnJ vaccine relative to the mRNA vaccines against the Delta variant ^31^.

There are several limitations to our study. First, breakthrough infections in this study were identified by testing of persons presenting to a tertiary hospital and clinic or as part of community-based testing by a commercial laboratory, so sampling bias may be present. This limitation is mitigated by our results showing similar variant distributions and viral load comparisons across two separate test cohorts. Second, the total number of vaccinated persons with breakthrough infections was relatively small at 125, of which clinical and epidemiologic metadata were only available for 39. Third, clinical data were obtained by retrospective medical chart review and thus may have had missing data or may have been subject to inaccurate reporting. Fourth, in the absence of contact tracing metadata, we were unable to assess transmission and secondary attack rates from vaccinated persons to exposed contacts.

In summary, our results reveal that selection pressure in a highly vaccinated community (>71% fully vaccinated as of early August 2021) favors vaccine breakthrough infections from antibody-resistant VOCs such as the Gamma ^12,32^and Delta ^12,13,21,25^ variants, and that high-titer symptomatic post-vaccination infections may be a key contributor to viral spread. Concerns have also been raised regarding waning immunity resulting in decreased effectiveness of the vaccine in preventing symptomatic infection over time ^33^. Combined with other potential factors such as relaxation of COVID-19 restrictions and complacency due to “pandemic fatigue”, these data may explain the recent steep rise in COVID-19 cases in San Francisco County (Figure 1) ^34^ and nationwide ^18^ in July-August 2021. Targeted booster vaccinations to increase protective neutralizing antibody levels against antibody-resistant variants ^35,39^, potentially guided by monitoring of immune correlates of vaccine efficacy^30^, will likely be needed in the near future to control viral spread in the community.

vaccinated individuals are more susceptible to COVID variant infections than unvaccinated individuals.” To table

## Data Availability

Data Availability
Assembled SARS-CoV-2 genomes in this study were uploaded to GISAID (accession numbers in Supplementary Table S2) and can be visualized in NextStrain. Viral genomes were also submitted to the National Center for Biotechnology Information (NCBI) GenBank database (accession numbers pending). Raw sequence data were submitted to the Sequence Read Archive (SRA) database. (BioProject accession number PRJNA722044 and umbrella BioProject accession number PRJNA171119).
Code Availability
FASTA files and scripting code for data analyses are available in a Zenodo data repository (https://doi.org/10.5281/zenodo.5207242).

https://doi.org/10.5281/zenodo.5207242

## Acknowledgments

We thank the UCSF Center for Advanced Technology core facility (Delsy Martinez and Tyler Miyasaki) for their efforts in high-throughput sequencing of viral cDNA libraries using the Illumina NovaSeq 6000 instrument.

## Funding

This work has been funded by US CDC Epidemiology and Laboratory Capacity (ELC) for Infectious Diseases Grant 6 NU50CK000539 to the California Department of Public Health (M-K.M., C.H., D.A.W.), the Innovative Genomics Institute (IGI) at UC Berkeley and UC San Francisco (C.Y.C.), National Institutes of Health grant R33AI129455 (C.Y.C.), US Centers for Disease Control and Prevention contract 75D30121C10991 (C.Y.C.), and Heluna Health/California Department of Public Health contract 5NU50CK000539 (C.Y.C.). The contents of this article are those of the author(s) and do not necessarily represent the official views of, nor an endorsement, by the CDC/HHS, San Francisco Department of Public Health, California Department of Public Health, or the U.S. Government.

## Author contributions

C.Y.C., S.P., S.T., and D.S. conceived and designed the study. C.Y.C, and V.S. coordinated the sequencing efforts and laboratory studies. V.S., M-K.M., A.S-G., E.T., B.W., D.W., C.W, Y.Z., N.B., K.R.R., D.R.G., X.D., and E.F. performed experiments. C.Y.C. and V.S. performed genome assembly and viral mutation analysis. C.Y.C., V.S., M.K.M, A.S-G., A.G., K.H., M.S., N.B., J.H.Jr., C.H. analyzed data. V.S., A.S.-G., A.G., E.T., N.B., A.Z., D.W., S.T., D.W., K.R.R., J.S., and S.M. collected samples. C.Y.C., and V.S. wrote the manuscript. C.Y.C. and V.S. prepared the figures. C.Y.C., V.S., M-K.M., N.B., B.W., D.W., C.W., Y.Z., X.D., J.H.Jr., C.H., D.W., and S.T. edited the manuscript. All authors read the manuscript and agree to its contents.

## Competing interests

C.Y.C. is the director of the UCSF-Abbott Viral Diagnostics and Discovery Center and receives research support from Abbott Laboratories, Inc. E.F. and J.H.,Jr. are employees and shareholders of Abbott Laboratories. E.T., A.Z., and S.T. are employees of Color Genomics. The other authors declare no competing interests.

## STAR Methods Text

### RESOURCE AVAILABILITY

#### Lead Contact

Further information and requests for resources and reagents should be directed to and will be fulfilled by the Lead Contact, Charles Chiu.

#### Materials Availability

This study did not generate any new reagents.

#### Data Availability

Assembled SARS-CoV-2 genomes in this study were uploaded to GISAID (accession numbers in Supplementary Table S2) and can be visualized in NextStrain. Viral genomes were also submitted to the National Center for Biotechnology Information (NCBI) GenBank database (accession numbers pending). Raw sequence data were submitted to the Sequence Read Archive (SRA) database. (BioProject accession number PRJNA722044 and umbrella BioProject accession number PRJNA171119).

#### Code Availability

FASTA files and scripting code for data analyses are available in a Zenodo data repository (https://doi.org/10.5281/zenodo.5207242).

## EXPERIMENTAL MODEL AND SUBJECT DETAILS

### METHOD DETAILS

#### Human Sample Collection and Ethics Statement

Remnant nasopharyngeal and/or oropharyngeal (NP/OP) samples and plasma samples from laboratory confirmed SARS-CoV-2 positive patients were retrieved from the UCSF Clinical Laboratories and stored in a biorepository until processed. Remnant samples were biobanked and retrospective medical chart review for relevant clinical and demographic metadata were performed under a waiver of consent and according to protocols approved by the UCSF Institutional Review Board (protocol number 10-01116, 11-05519).

De-identified samples from community COVID-19 testing were obtained from Color Genomics Laboratory as part of a research collaboration. Vaccine breakthrough data corresponding to the de-identified samples from Color Genomics were obtained from the San Francisco Department of Public Health. As these data was obtained as part of SFDPH COVID-19 surveillance, institutional review board approval was not required.

#### Viral Whole-Genome Sequencing

For primary nasopharyngeal and/or oropharyngeal swab samples from UCSF hospitals and clinics, remnant samples collected in UTM/VTM were diluted with DNA/RNA shield (Zymo Research, # R1100-250) in a 1:1 ratio (100 ul primary sample + 100 ul shield). The Omega BioTek MagBind Viral DNA/RNA Kit (Omega Biotek, # M6246-03) and the KingFisherTM Flex Purification System with a 96 deep-well head (ThermoFisher, 5400630) were then used for viral RNA extraction. For mid-turbinate nasal swab samples sent to Color Genomics for commercial laboratory testing, dry swabs were collected and transported to the laboratory with no added media. At the laboratory, the swabs were resuspended in 1.3 mL of lysis buffer and RNA was extracted using the Chemagic 360 system (Perkin-Elmer). Remnant RNA was then aliquoted for viral whole-genome sequencing.

Extracted RNA was reverse transcribed to complementary DNA and tiling multiplexed amplicon PCR was performed using SARS-CoV-2 primers version 3 according to a published protocol^37^. Adapter ligation was performed using the NEBNext Ultra II DNA Library Prep Kit for Illumina (New England Biolabs, # E7645L). Libraries were barcoded using NEBNext Multiplex Oligos for Illumina (96 unique dual-index primer pairs) (New England Biolabs, # E6440L) and purified with AMPure XP (Beckman-Coulter, #. Amplicon libraries were then sequenced on either Illumina NextSeq 550 or Novaseq 6000 as 1×300 single-end reads (300 cycles).

#### Genome Assembly, Variant Calling and Viral Mutation Analysis

SARS-CoV-2 viral genome reads were assembled and variants were identified using an in-house bioinformatics pipeline as previously described^38^. BCL files generated by Illumina sequencers (NextSeq 550 or NovaSeq 6000) were simultaneously demultiplexed and converted to FASTQ files. Raw FASTQ files were first screened for SARS-CoV-2 sequences using BLASTn (BLAST+ package 2.9.0) alignment against viral reference genome NC_045512. Reads containing adapters, the ARTIC primer sequences, and low-quality reads were filtered using BBDuk (version 38.87), and then mapped to the NC_045512 reference genome using BBMap (version 38.87). Variants were called with CallVariants and a depth cutoff of 5 was used to generate the final assembly. Pangolin software (version 3.0.2) was used to identify the lineage ^36^. Using a custom in-house script, consensus FASTA files generated by the genome assembly pipeline were scanned to confirm the presence/absence of resistance-associated (L452R, L452Q, E484K, and/or F490S), and infectivity-associated (L452R/Q, F490S, and/or N501T/Y) mutations. Only genomes with defined lineages were included in this analysis.

#### RT-PCR and Viral Load Analysis

The TaqPath COVID-19 Combo kit (ThermoFisher) was used to determine cycle threshold (Ct) values. This multiplex Real-time RT-PCR assay detects the nucleoprotein (N) gene, spike (S) gene, and orf1ab genes. For simplicity, only the N gene Ct value was used for quantitative analysis of RNA viral loads in this study.

#### Antibody Assays

SARS-CoV-2-specific antibodies were determined using the Abbott ARCHITECT SARS-CoV-2 IgG (N-based), AdviseDx SARS-CoV-2 IgM (spike receptor-binding domain (RBD)-based), and AdviseDx SARS-CoV-2 IgG II (spike RBD-based) tests according to the manufacturer’s specifications.

#### CPE endpoint neutralization assays using a VOC lineage virus

CPE endpoint neutralization assays were done following the limiting dilution model (Wang et al., 2005) and using P1 stocks of D614G, B.1.1.7, B.1.617.2, B.1.429, B.1.351 and P.1 lineages. Convalescent patient plasma was diluted 1:10 and heat inactivated at 56C for 30 min. Serial 2-fold dilutions of plasma were made in BSA-PBS. Plasma dilutions were mixed with 100 TCID50 of each virus diluted in BSA-PBS at a 1:1 ratio (160ul plasma dilution and 160ul virus input) and incubated for 1 hour at 37C. Final plasma dilutions in plasma-virus mixture ranged from 1:100 to 1:12800. 100 uL of the plasma-virus mixtures were inoculated on confluent monolayer of Vero-81 cells in 96-well plates in triplicate and incubated at 37C with 5% CO2 incubator. After incubation 150 uL of MEM containing 5% FCS was added to the wells and plates were incubated at 37C with 5% CO2 until consistent CPE was seen in virus control (no neutralizing plasma added) wells. Positive and negative controls were included as well as cell control wells and a viral back titration to verify TCID50 viral input. Individual wells were scored for CPE as having a binary outcome of ‘infection’’ or ‘no infection’ and the IC50 was calculated using the Spearman-Karber method. All steps were done in a Biosafety Level 3 lab using approved protocols.

## STATISTICAL ANALYSES

Statistical analysis was performed using Python scipy package (version 1.5.2) and rstatix package (version 0.7.0) in R (version 4.0.3). For comparisons of the mean cycle threshold (Ct) values, significance testing was done using Welch’s t-test as implemented in Python (version 3.7.10). Fisher’s exact test method was used to assess the association of demographics and clinical variables with vaccination status. Box-and-whisker and swarm plots were generated using Python matplotlib (version 3.3.2) and seaborn (version 0.11.0) packages. All statistical tests were conducted as two-sided at the 0.05 significance level.

## Supplementary Figures and Tables

**Supplementary Table 1.** Vaccine received and number of days from completion of vaccine to COVID-19 infection for 39 confirmed vaccine breakthrough cases.

| Patient | # of days from completion of vaccine doses to positive COVID-19 diagnostic test | vaccine type |
| --- | --- | --- |
| 1 | 37 | Pfizer |
| 2 | 49 | Pfizer |
| 3 | 48 | Pfizer |
| 4 | 23 | Pfizer |
| 5 | 44 | Pfizer |
| 6 | 17 | Pfizer |
| 7 | 59 | Pfizer |
| 8 | 42 | Moderna |
| 9 | 42 | Moderna |
| 10 | 20 | not specified |
| 11 | 94 | Pfizer |
| 12 | 83 | Pfizer |
| 13 | 27 | J&J |
| 14 | 84 | Pfizer |
| 15 | 30 | J&J |
| 16 | 78 | Moderna |
| 17 | 117 | Pfizer |
| 18 | 18 | J&J |
| 19 | 69 | Pfizer |
| 20 | 81 | Moderna |
| 21 | 129 | Pfizer |
| 22 | 116 | Pfizer |
| 23 | 47 | J&J |
| 24 | 31 | Moderna |
| 25 | 118 | Moderna |
| 26 | 140 | Pfizer |
| 27 | 58 | Moderna |
| 28 | not specified | not specified |
| 29 | 102 | Pfizer |
| 30 | 133 | Moderna |
| 31 | 36 | not specified |
| 32 | 123 | not specified |
| 33 | 15 | Moderna |
| 34 | 104 | Pfizer |
| 35 | 114 | Pfizer |
| 36 | 131 | Moderna |
| 37 | 81 | Pfizer |
| 38 | 112 | Pfizer |
| 39 | 127 | Moderna |

**Supplementary Table 2. Metadata for the 945 clinical samples with identified lineages and/or mutations included in this study**.

(separate file in Excel format, “SupplementaryTable2.xlsx”)

**Figure S1.**
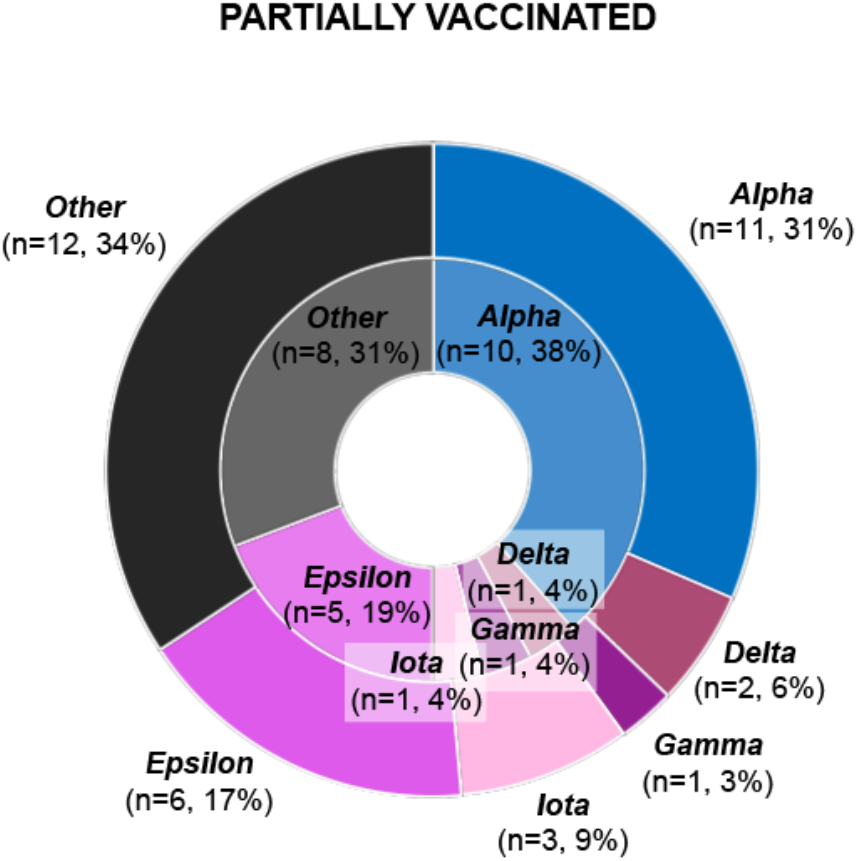
Lineage distribution and proportion of mutations in partially vaccinated cases from UCSF Hospitals and Clinics. The inner circles represent the immunocompetent cases, and the outer circles include both immunocompetent and immunocompromised individuals.

**Figure S2.**
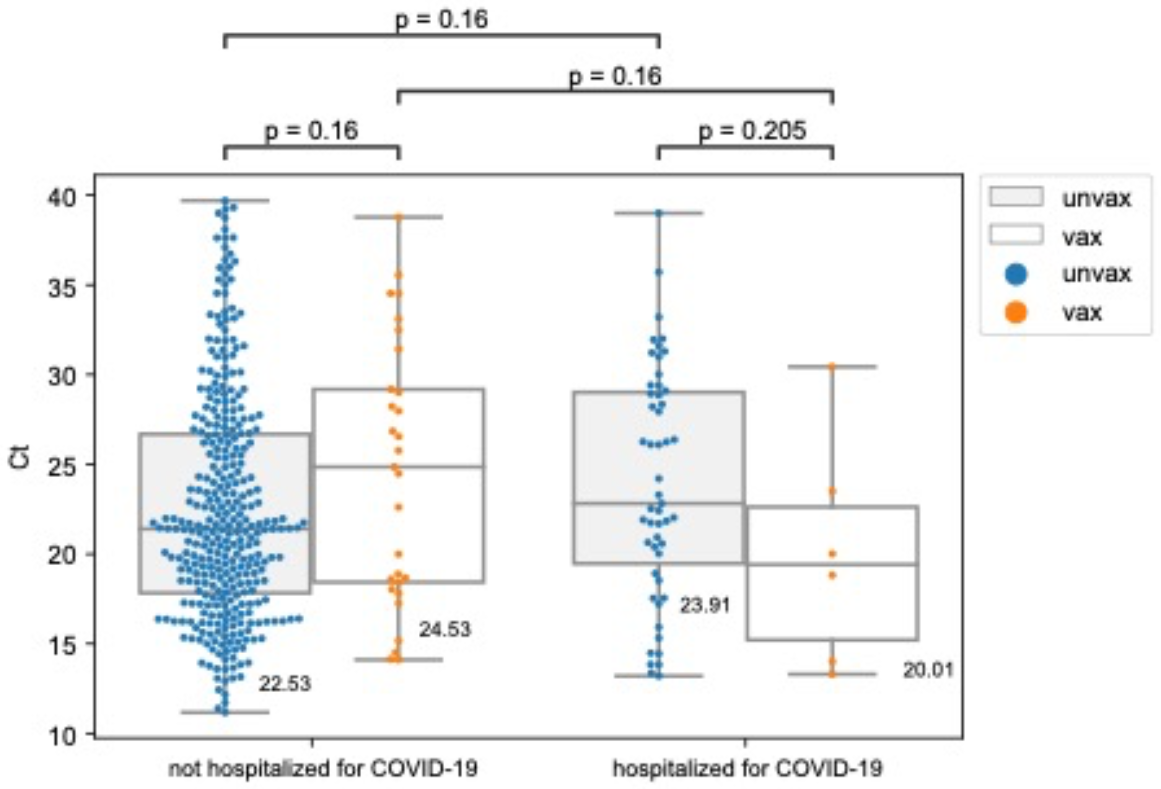
Comparison of viral loads between vaccinated and unvaccinated cases based on hospitalization status. Grouped box-and-whisker plots and swarm plots showing the differences in mean cycle threshold (Ct) values between vaccinated and unvaccinated cases overall given hospitalization (due to COVID-19) status. There were no significant differences in viral loads (inversely proportional to the Ct value) in the pairwise comparisons.

## Notes

### Author Declarations

Remnant nasopharyngeal and/or oropharyngeal (NP/OP) samples and plasma samples from laboratory confirmed SARS-CoV-2 positive patients were retrieved from the UCSF Clinical Laboratories and stored in a biorepository until processed. Remnant samples were biobanked and retrospective medical chart review for relevant clinical and demographic metadata were performed under a waiver of consent and according to protocols approved by the UCSF Institutional Review Board (protocol number 10-01116, 11-05519). De-identified samples from community COVID-19 testing were obtained from Color Genomics Laboratory as part of a research collaboration. Vaccine breakthrough data corresponding to the de-identified samples from Color Genomics were obtained from the San Francisco Department of Public Health. Approval for sequencing and analysis of these de-identified samples and metadata was obtained from the UCSF Institutional Review Board (protocol number 11-05519).

### Summary of Updates

author list updated and disclaimer added to the Funding

